# Using Vision and Large Language Models for Psychological Intervention Competency Assessment: A Pilot Study of Agreement with Clinician Ratings

**DOI:** 10.64898/2026.11.02.26367108

**Authors:** Yinuo Liu, William Leever, Maddie Stanco, Zoe Shook, Prisilla Addico, Sterling Gleeson, Florencia Nugroho, Grace Varney, Eric Youngstrom, Emre Sezgin

## Abstract

**Introduction:** Supervision in psychological intervention training is limited by the time needed to evaluate simulated sessions. We tested a Vision Language Model (VLM) and Large Language Model (LLM) pipeline that assesses competency from verbal and nonverbal communication.

**Method:** Gemini-3.1-Pro was used to annotate behaviors in 25 simulated therapy videos and incorporate them into transcripts, and ChatGPT-6.1-Sol rated them against the ENACT scale. Two clinicians verified VLM-identified behaviors and four independently rated each video. Agreement was estimated with Gwet’s AC1 and ICC.

**Results:** Of 1,558 VLM-identified behaviors, 70% were confirmed by both clinicians and 93% by at least one. AI-clinician agreement on therapeutic behavior attributes (AC1=0.73, 95% CI 0.70-0.76) approached between-clinician agreement (0.75, 0.72-0.78). AI-clinician ICCs on therapeutic domain levels (median 0.57) were modestly lower than between-clinician ICCs (0.67).

**Discussion:** The pipeline agreed with clinicians nearly as well as clinicians agreed with each other and shows promise as a supplementary evaluator in therapist training.

## Introduction

The prevalence of depression among adolescents and adults in the United States rose from 8.2% in 2013-2014 to 13.1% in 2021-2023,[1] and diagnosed anxiety among children increased by 29% between 2016 and 2020,[2] raising demand for evidence-based psychological interventions that the clinical workforce has not met. About 137 million Americans live in federally designated mental health professional shortage areas, with a projected shortfall of 21,480 mental health counselors in 2026.[3] Scaling up psychological interventions is one response to this gap, but expanding this workforce depends on supervised training, in which a supervisor watches each simulated session, rates it against a structured rubric, and documents feedback, so the supply of supervisors and the time per review constrain how many trainees can be trained and how often they receive feedback.

Global efforts in task sharing of psychological interventions offer a model for scaling provider training and competency assessment. The World Health Organization (WHO) has developed brief, scalable psychological interventions, which use elements of cognitive behavioral therapy for communities with limited specialist providers.[4] The accompanying implementation manual guides the integration of these interventions into existing services and treats workforce preparation as a core step in expanding access to care. As the non-specialist workforce expands, competency-based training becomes essential to the quality of care. New providers must demonstrate foundational helping skills, such as rapport building and nonverbal communication, as well as intervention-specific skills, such as problem solving or behavioral activation. The training needs of non-specialist providers are easily underestimated,[5] and the supervision they require falls on specialists who already carry high caseloads, so reducing the supervision burden is a condition for scaling this model of care.

Automating the evaluation with artificial intelligence (AI) could relieve this training and supervision bottleneck. Existing AI approaches to rating therapist competence or fidelity have worked from transcripts alone,[6–8] which carry only conversational content. Eye contact, facial expressions, gestures, posture, and tone of voice are central to rapport building [9] and carry important assessment cues for various competency domains, so a rating built from dialogue alone cannot provide the evidence those domains require.

Recent developments in Vision Language Models (VLMs) and Large Language Models (LLMs) offer a way to address this challenge. VLMs accept images and audio and can describe visual and vocal behaviors alongside the spoken content, converting a video into a text record of verbal and nonverbal communication. LLMs can evaluate text consistently against a pre-specified rubric, applying the same criteria to every session. Separating perception from judgment also makes the assessment auditable, since the behavior-annotated transcript is a record that a supervisor can check against the video and to which each rating can be traced. We therefore tested a VLM+LLM pipeline on simulated therapy sessions, asking whether the VLM captures verbal and nonverbal communication accurately and whether LLM ratings of the resulting transcripts agree with clinician ratings.

## Methods

### Material

We compiled 25 publicly available videos of simulated therapy sessions from YouTube, 2 to 32 minutes long, comprising 11 intake sessions, 4 motivational interviewing sessions, and 10 demonstrations of flawed counseling skills (selection criteria in Table S1, video details in Table S2). The flawed-skill videos were purposively selected so that unhelpful or potentially harmful behaviors would be represented.

### Rubric

We used the ENhancing Assessment of Common Therapeutic factors (ENACT) (English, in-person, v1.0), which was developed for training and supervision of psychological treatments and has shown acceptable validity and inter-rater reliability.[10] For each therapeutic domain, ENACT lists binary behavior attributes grouped as unhelpful or potentially harmful behaviors, basic helping skills, and advanced helping skills. Each domain also receives a level from 1 to 4, with higher levels indicating stronger competency. Clinicians assigned domain levels by holistic judgment, whereas the LLM derived each level from the attributes it marked present by applying explicit scoring rules adapted based on clinician practices (Table S3). We excluded Domain 7 (Assessment of Harm and Developing Response Plan) because harm management requires dedicated sessions and could not be assessed in our material.

### Evaluators

Six Qualified Behavioral Health Specialists employed at Nationwide Children’s Hospital volunteered as evaluators. Evaluators had between 16 and 22 months of clinical experience and used the ENACT weekly in clinical supervision. Two evaluators verified VLM-identified behaviors, and the other four rated the videos against the ENACT.

### Model

We used the VLM Gemini-3.1-Pro via API to capture verbal and nonverbal communication in simulated therapy videos. Each video or selected clip was input at one frame per second with its audio track, title, and metadata. The VLM was prompted to transcribe the dialogue and to annotate timestamped behaviors, labeling each as descriptive (directly observable) or interpretative (inferring emotion or interactional function).

We used the LLM ChatGPT-6.1-Sol via API to rate the behavior-annotated transcripts against the ENACT. For each domain, the LLM marked each behavior attribute as present or absent, assigned a domain level based on specified rules, and provided scoring rationale by citing timestamped evidence from the transcript. Attributes and levels could be rated Not Assessable (NA) when the transcript offered no opportunity to demonstrate the skill. Model access details and full prompts are in Table S3.

### Procedure

In the pipeline (Figure 1), the VLM processed each video into a behavior-annotated transcript, the LLM rated each transcript against the ENACT, and two human evaluation tasks proceeded in parallel. Two evaluators judged whether each VLM-identified behavior was present in the original video, and four evaluators independently rated every video against the ENACT, blinded to the transcripts and AI ratings. All raters evaluated the videos in the same order and could view video titles and metadata.

**Figure 1.**
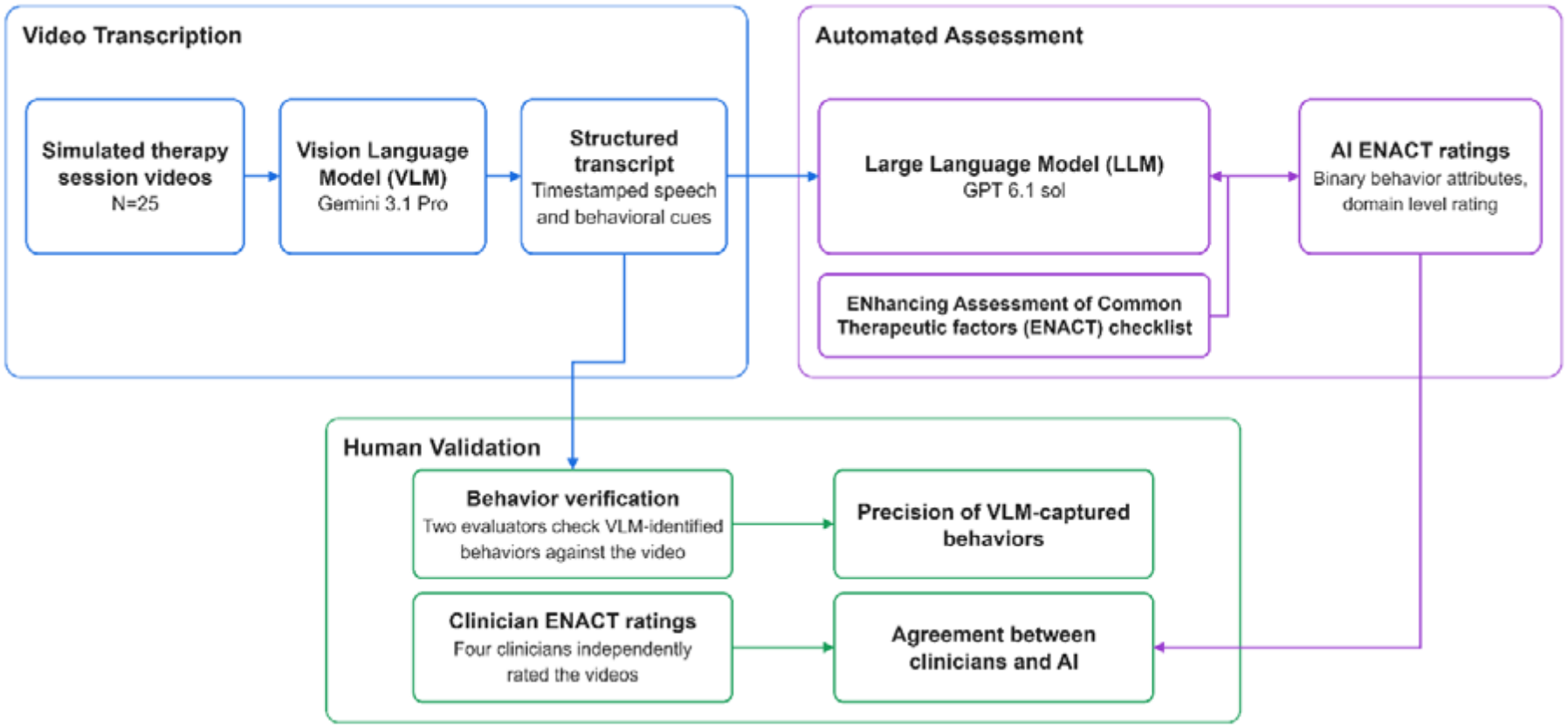
Overview of the VLM+LLM assessment pipeline for psychological intervention competency.

### Analysis

All analyses were conducted in R 4.6.0 with the *irr* and *irrCAC* packages. [11–13]

#### VLM behavior precision

Precision was defined as the percentage of VLM-identified behaviors confirmed by the two human evaluators. We report it under a strict criterion (both evaluators confirmed) and a lenient criterion (at least one evaluator confirmed). Agreement between the two evaluators was summarized with Gwet’s AC1,[14] with thresholds defined as slight (<0.2), fair (0.2-0.4), moderate (0.4-0.6), substantial (0.6-0.8), and excellent (0.8-1.0).[15]

#### Behavior rating agreement

Between-clinician and AI-clinician agreement on ENACT binary behavior attributes was calculated with Gwet’s AC1, overall and separately for harmful behaviors (only in videos selected to demonstrate them), basic skills, and advanced skills. AC1 was computed on video-attribute ratings pooled across videos, using the multi-rater form across four clinicians for between-clinician agreement and the four AI-clinician pairs stacked together for AI-clinician agreement. Because the clinician checklist does not include a not-assessable option, behaviors the AI rated as NA were treated as absent.

#### Domain level agreement

Agreement on the 1–4 ENACT domain level was estimated with a one-way, single-measure intraclass correlation coefficient (ICC(1)), interpreted with the same thresholds as AC1.[15] Blank clinician levels and AI NA ratings were treated as not assessable. ICC was computed on all rater pairs stacked together, keeping only pairs in which both raters gave a valid level. Primary analyses only included domains with valid clinician and AI ratings in more than 20 videos.

#### Confidence intervals

95% CIs for AC1 and ICC were bias-corrected and accelerated (BCa) bootstrap intervals (2,000 resamples of videos), which adjust for skewness in bounded agreement coefficients.[16]

## Results

### VLM Behavior Frequency

The VLM identified 1,558 behaviors across the 25 videos, most of them descriptive, with gestures, head movements, and visual attention accounting for most annotations and vocal cues and interpersonal distance rarely captured (Table 1). The number of behaviors per video ranged from 20 to 146, and behavior density was higher in short clips demonstrating unhelpful behaviors than in full-length intake sessions (Table S4).

**Table 1.** Frequency of VLM-identified behaviors across categories.

| **Behavior Category** | **Frequency** | **Percentage** |
|---|---|---|
| Total | 1558 | 100.0% |
| Interpretative | 179 | 11.5% |
| Descriptive | 1379 | 88.5% |
| Gesture | 657 | 42.2% |
| Head Movement | 221 | 14.2% |
| Visual Attention | 186 | 11.9% |
| Body Posture | 100 | 6.4% |
| Facial Expression | 99 | 6.4% |
| Eye Contact | 89 | 5.7% |
| Vocal Cue | 26 | 1.7% |
| Interpersonal Distance | 1 | 0.1% |

### VLM Behavior Precision

Overall, 70% of VLM-identified behaviors were confirmed by both evaluators (strict criterion) and 93% by at least one (lenient criterion). Interpretative behaviors had higher precision than descriptive behaviors under both criteria (strict 78.2% vs. 68.5%, lenient 99.4% vs. 91.7%). Among descriptive categories, strict precision was highest for eye contact (88.8%) and lowest for body posture (37.0%), and lenient precision was at least 85.0% in every category (Table 2).

**Table 2.** Precision of VLM-identified behaviors along with rater agreement.

| **Behavior Category** | **Strict precision** | **Lenient precision** |
|---|---|---|
| Total | 70% | 93% |
| Interpretative | 78% | 99% |
| Descriptive | 69% | 92% |
| Gesture | 67% | 88% |
| Head Movement | 76% | 99% |
| Visual Attention | 74% | 93% |
| Body Posture | 37% | 85% |
| Facial Expression | 66% | 96% |
| Eye Contact | 89% | 99% |
| Vocal Cue | 73% | 100% |
| Interpersonal Distance | 100% | 100% |
| **Rater Agreement** | **Gwet's AC1** | **95% CI** |
| Total | 0.67 | 0.63-0.71 |
| Interpretative | 0.74 | 0.64-0.83 |
| Descriptive | 0.66 | 0.62-0.70 |

The two evaluators showed substantial agreement in confirming VLM-identified behaviors (overall AC1 0.67, 95% CI 0.63-0.71). Agreement was similar for interpretative behaviors (0.74, 0.64-0.83) and descriptive behaviors (0.66, 0.62-0.70), with overlapping confidence intervals. The difference between strict and lenient precision therefore reflects disagreement between evaluators about what the video shows as much as it reflects VLM error.

### Behavior Rating Agreement

The LLM completed each evaluation in a median of 64s (range: 53-84s) at a cost of $0.07-$0.09 per video (Table S4). Across all binary ENACT behavior attributes, AI-clinician agreement was substantial (AC1 0.73, 95% CI 0.70-0.76) and close to between-clinician agreement (0.75, 0.72-0.78) (Figure 2). The two comparisons were also comparable within behavior subcategories. For unhelpful or potentially harmful behaviors, AI-clinician agreement (0.65, 0.54-0.76) was slightly higher than between-clinician agreement (0.59, 0.49-0.71). Agreement was substantial for advanced helping skills (between-clinician 0.73, 0.60-0.84, AI-clinician 0.73, 0.61-0.82) and moderate for basic helping skills (between-clinician 0.59, 0.54-0.68, AI-clinician 0.53, 0.47-0.61). In every category, AI-clinician agreement was within 0.07 of between-clinician agreement, and the bootstrapped confidence intervals of the two comparisons overlapped.

**Figure 2.**
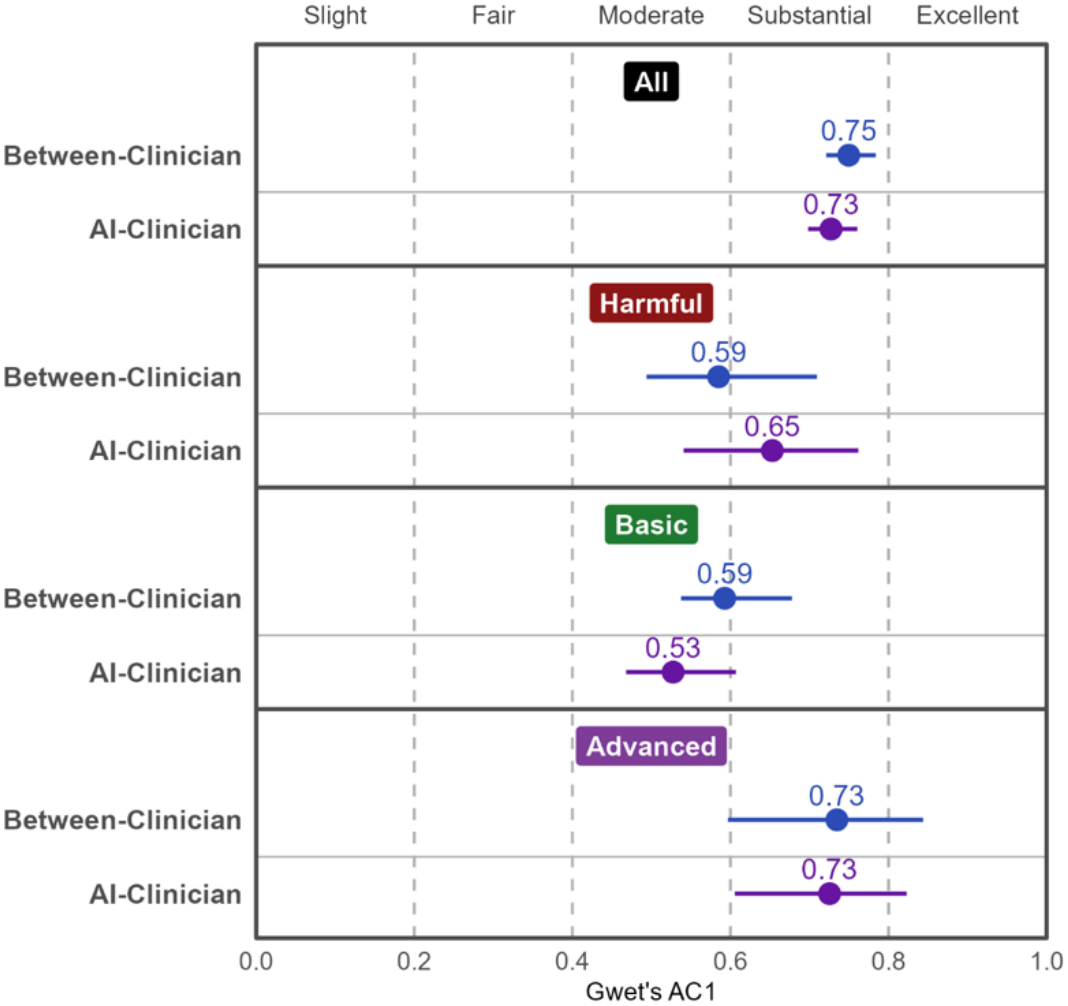
Between-clinician and AI-clinician agreement calculated with Gwet’s AC1 for ENACT binary behavior attributes, overall and separately for unhelpful or potentially harmful behaviors, basic helping skills, and advanced helping skills.

### Domain Level Agreement

The AI rated two domains not assessable more often than clinicians did, providing valid levels in 13 videos for Confidentiality and 14 for Rapport Building. These classifications were concentrated in the motivational interviewing and flawed-skill videos, which begin mid-session, and they follow from the LLM prompt, which instructed the model to treat introductions and a confidentiality briefing as not assessable when a session was not a first meeting. The AI’s NA ratings therefore follow a consistent, prompt-defined rule. Clinicians received no equivalent instruction, and their blank levels varied considerably across individuals, with one clinician accounting for most of them. These blank levels more likely reflect individual practice than a shared criterion (Table S5).

In other ENACT domains, between-clinician ICCs ranged from moderate to excellent (0.43–0.93) and were substantial in 8 of 12 domains (Figure 3). AI-clinician ICCs ranged from moderate to substantial (0.46–0.70). Overall, AI-clinician agreement was modestly lower than between-clinician agreement (median ICC 0.57 vs. 0.67). The largest discrepancy was observed for Verbal Communication, where between-clinician agreement was excellent (0.93), the highest of any domain, and AI-clinician agreement was only moderate (0.55). In Family Involvement and Coping Mechanisms, AI agreed better with clinicians than clinicians agreed with each other (0.56 vs. 0.43, and 0.60 vs. 0.57). Table S6 presents ICCs with 95% CIs for all domains assessed.

**Figure 3.**
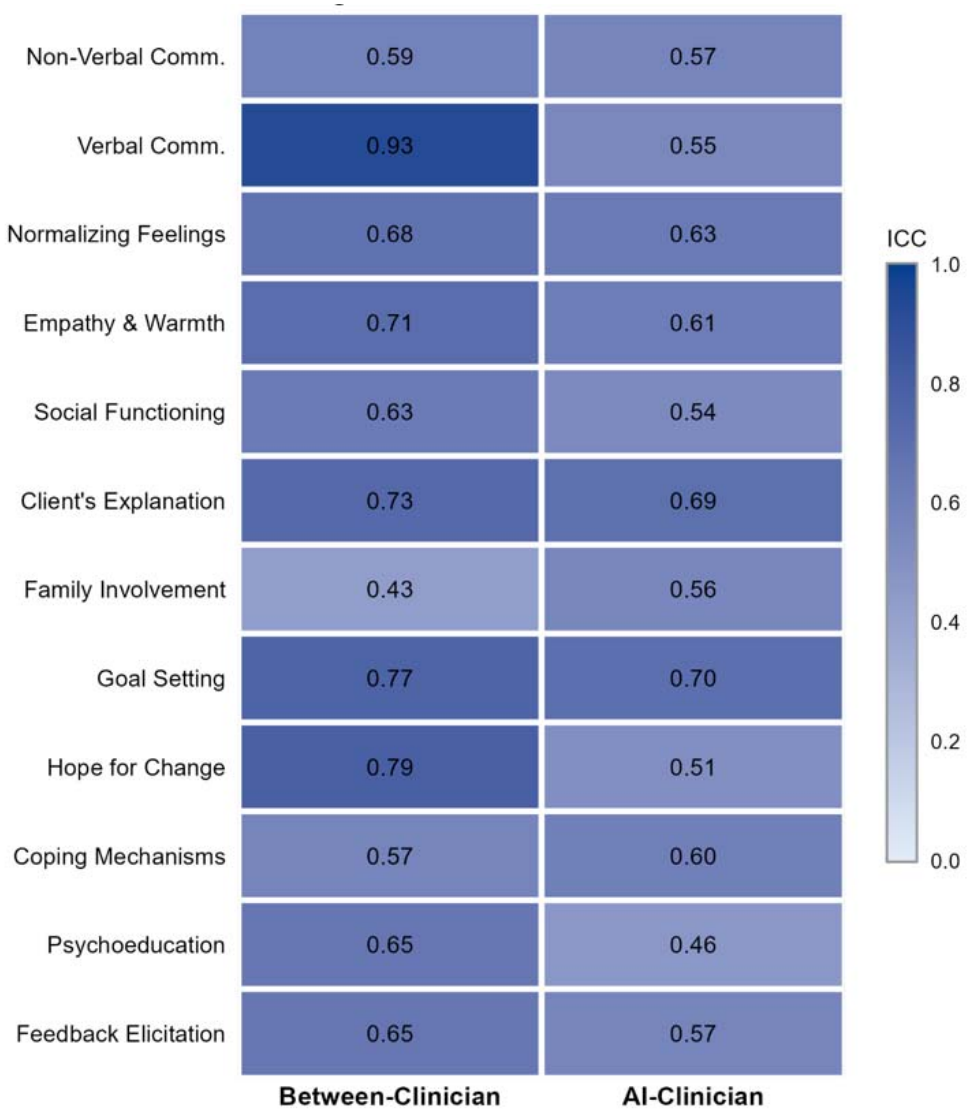
Between-clinician and AI-clinician agreement on ENACT therapeutic domains calculated with Intraclass Correlation Coefficients (ICC).

## Discussion

This pilot study evaluated a pipeline combining a Vision Language Model and a Large Language Model to assess psychological intervention competency in simulated therapy sessions. Most VLM-identified behaviors were verified in the original videos, and the AI ENACT ratings agreed substantially with clinicians on binary behavior attributes and moderately to substantially on domain levels, comparable to or modestly below between-clinician agreement.

These results should be read against the variability of the human benchmark. Clinicians did not align perfectly in behavior confirmation (AC1=0.67), attribute presence (AC1=0.75), or domain-level rating (ICC 0.43–0.93), indicating they interpret video content and ENACT criteria differently. Because no single clinician provides a definitive reference standard, consistency among clinicians sets a practical upper bound on the agreement any rater, human or automated, can reach.[15] The VLM+LLM pipeline came close to this agreement upper bound on behavior attributes and fell modestly short on domain levels. It can therefore be positioned as an additional rater whose judgments fall within the range of disagreement already present among clinicians, rather than as a substitute for them.

The pipeline applies the same criteria to every session without fatigue, requires little manual effort beyond recording and uploading sessions, and produces standardized ratings with timestamped behavior-annotated transcripts that are easier to store, search, and audit than raw video. Processing and rating each video took under 6 minutes and cost less than $1, for a total of about $12 across 247 minutes of footage (Table S4). Because each rating is supported by time-stamped evidence from the transcript, supervisors can verify ratings quickly and focus their review where human judgment is most needed.

These findings have practical implications for how the pipeline can be integrated into clinical training and supervision. Competence in psychological interventions develops through repeated practice with specific and timely feedback,[17] and the supervisors who can provide such feedback are a limited resource in most training settings.[18] An automated evaluator that returns rubric-anchored and evidence-linked feedback after every session can potentially change the frequency of feedback that trainees receive (e.g., feedback on each simulated session rather than on a sample of sessions), which may support skill acquisition over the course of training. However, the use of automated ratings should remain formative at this stage. Observer-rated competence has shown only modest associations with client outcomes,[19] which indicates that an automated rating is an indicator for training instead of a warrant of quality.

Therefore, its use in certification or employment decisions should be deferred until its relationship with outcomes has been examined. Transcript-based fidelity feedback has been found feasible and acceptable to therapists and community mental health supervisors,[20] and a similar implementation path can be expected for a multimodal evaluator. Here, feedback could be delivered to the trainee after each session, and supervisor review is reserved for flagged sessions. Such an approach can potentially place clinician expertise where disagreement is most informative, instead of requiring supervisor review across every session.

The results also point to where the pipeline should change. The VLM annotated behavior categories unevenly, body posture had the lowest precision, and vocal cues accounted for only 26 of 1,558 annotations even though the prompt named tone, pace, pauses, and silence as a category to capture, so the transcript carried almost no indication about how a message was verbally conveyed or person self-expressed. This sparsity is a plausible source of the largest divergence in domain levels, since clinicians agreed almost perfectly on Verbal Communication (ICC 0.93) while hearing the sessions, whereas the LLM rated it from text with little paralinguistic evidence (0.55). Because prompting alone did not elicit paralinguistic annotation, a separate audio pass or acoustic features extracted independently of the visual stream may be needed, and a behavior list derived from the ENACT attributes rather than improvised for this pilot would align annotation with the cues the rubric scores. In general, clinicians agreed more with each other than with the AI on domain levels, which may reflect a shared understanding of the criteria, built through clinical training, that the prompt does not capture.

Several constraints bound these inferences. The sample was small, covered three session types, and consisted of public videos of simulated sessions that varied in length, resolution, and recording angle, so generalization to recorded clinical sessions is untested. Although clinicians rated without access to transcripts or AI ratings, neither they nor the models were blinded to video titles and metadata, and because several titles announce flawed practice, both could infer the intended quality of a session, which may have inflated agreement on harmful behaviors in particular. The evaluators were early-career clinicians who use ENACT routinely, so the benchmark reflects trained routine raters rather than experienced supervisors. We assessed precision but not recall of VLM-identified behaviors, and each video was processed once at a sampling temperature of 1, so missed behaviors remain unquantified and the stability of AI ratings across runs and model versions is unknown. The comparison is one of agreement rather than accuracy, since no reference standard for competency exists beyond clinician judgment, and VLM identification of cues such as eye contact and interpersonal distance may carry cultural assumptions that these sessions did not test.

Next steps are to refine the prompts, assess recall and repeat-run stability, test alternative models, and scale to standardized simulated sessions recorded in a hospital training program, where any extension to real sessions would require privacy-preserving, on-premise model deployment.

In conclusion, an automated VLM-LLM pipeline produced competency ratings that agreed with clinicians nearly as well as clinicians agreed with each other, and could serve as an additional evaluator in therapist training once its recall and its performance on recorded clinical sessions have been established. This could help expand access to psychological interventions as the workforce scales up using task sharing approaches.

## Data Availability

All data produced in the present work are contained in the manuscript.

## Ethics

This study is based on publicly available videos without human subjects involved.

## Data Availability

All available data has been presented with the publication.

## Authors’ Contributions

Author roles were classified using the Contributor Role Taxonomy (CRediT; https://credit.niso.org/) as follows:

Yinuo Liu: data curation, investigation, formal analysis, and writing – original draft.

William Leever: conceptualization, writing – original draft, review & editing.

Maddie Stanco: validation, review & editing

Zoe Shook: validation, review & editing

Prisilla Addico: validation, review & editing

Sterling Gleeson: validation, review & editing

Florencia Nugroho: validation, review & editing

Grace Varney: validation, review & editing

Eric A. Youngstrom: supervision, review & editing.

Emre Sezgin: conceptualization, supervision, methodology, writing – original draft, review & editing

## References

1 Brody DJ, Hughes JP. Depression prevalence in adolescents and adults: United States, August 2021–August 2023. 2025.

2 Lebrun-Harris LA, Ghandour RM, Kogan MD, et al. Five-year trends in US children’s health and well-being, 2016-2020. JAMA pediatrics. 2022;176:e220056.

3 Health Resources and Services Administration. Health Workforce Projections. Bureau of Health Workforce. https://data.hrsa.gov/topics/health-workforce/nchwa/workforce-projections (accessed 20 October 2026)

4 World Health Organization. Psychological interventions implementation manual: Integrating evidence-based psychological interventions into existing services. World Health Organization 2024.

5 Kohrt BA, Mutamba BB, Luitel NP, et al. How competent are non-specialists trained to integrate mental health services in primary care? Global health perspectives from Uganda, Liberia, and Nepal. International Review of Psychiatry. 2018;30:182–98.

6 Lilan S, Mochama B, Osborn T, et al. Automated Fidelity Monitoring of Lay-Delivered Mental Health Interventions Using Large Language Models: Development and Pilot Validation of shamiriAI in Kenya. JMIR AI. 2026;5:e95063.

7 Flemotomos N, Martinez VR, Chen Z, et al. Automated quality assessment of cognitive behavioral therapy sessions through highly contextualized language representations. PloS one. 2021;16:e0258639.

8 Dillon C, Evans SW, Owens JS. Assessing the Interrater Agreement of Artificial Intelligence Transcript Analysis for Use in Consultation Training. Journal of Educational and Psychological Consultation. 2025;1–16. doi: 10.1080/10474412.2025.2529271

9 Ramseyer F, Tschacher W. Nonverbal synchrony in psychotherapy: coordinated body movement reflects relationship quality and outcome. Journal of consulting and clinical psychology. 2011;79:284.

10 Kohrt BA, Jordans MJD, Rai S, et al. Therapist competence in global mental health: Development of the ENhancing Assessment of Common Therapeutic factors (ENACT) rating scale. Behaviour Research and Therapy. 2015;69:11–21. doi: 10.1016/j.brat.2015.03.009

11 R Core Team. R: A Language and Environment for Statistical Computing. 2026.

12 Gamer M, Lemon J, Fellows I, et al. irr: Various Coefficients of Interrater Reliability and Agreement. 2005;0.85.

13 Gwet KL. irrCAC: Computing the Extent of Agreement among Raters with Chance-Corrected Agreement Coefficient (CAC). 2019;1.4.

14 Gwet KL. Computing inter□rater reliability and its variance in the presence of high agreement. British Journal of Mathematical and Statistical Psychology. 2008;61:29–48.

15 Landis JR, Koch GG. The measurement of observer agreement for categorical data. biometrics. 1977;159–74.

16 Efron B. Better bootstrap confidence intervals. Journal of the American statistical Association. 1987;82:171–85.

17 Chow DL, Miller SD, Seidel JA, et al. The role of deliberate practice in the development of highly effective psychotherapists. Psychotherapy. 2015;52:337.

18 Elnasseh A, Mehta VS, Manolova G, et al. Perspectives on competency-based feedback for training non-specialists to deliver psychological interventions: multi-site qualitative study of the EQUIP competency-based approach. BJPsych open. 2024;10:e125.

19 Webb CA, DeRubeis RJ, Barber JP. Therapist adherence/competence and treatment outcome: A meta-analytic review. Journal of consulting and clinical psychology. 2010;78:200.

20 Imel ZE, Pace BT, Soma CS, et al. Design feasibility of an automated, machine-learning based feedback system for motivational interviewing. Psychotherapy. 2019;56:318.

