## Supplemental materials for "Using Vision and Large Language Models for Psychological Intervention Competency Assessment: A Pilot Study of Agreement with Clinician Ratings"

Supplemental Table 1. Simulated therapy session selection criteria

| | |
|---|---|
| Platform | YouTube (https://www.youtube.com) |
| Search terms | "therapist role-play" "counseling role-play" "mental health role-play" "mock therapy session" "intake session role-play," "bad examples of therapist skills" "motivational interviewing" |
| Inclusion Criteria | 1. The video depicts a simulated (role-played) therapy or counseling session, as indicated by the title, description, or content.<br>2. The session is conducted in person, with the therapist and client physically present in the same room.<br>3. The session is dyadic, involving one therapist and one client.<br>4. The session is conducted in English.<br>5. Dialogue is audible and intelligible to permit transcription.<br>6. The video is publicly accessible without age restriction or a subscription. |
| Exclusion Criteria | 1. Telehealth or videoconference format.<br>2. Video resolution lower than 360p.<br>3. The therapist or client is outside the frame for more than 10% of the session duration.<br>4. Narration, instructor commentary, or on-screen text overlaps the interaction for more than 10% of its duration.<br>5. Group, couple, family, or child/adolescent sessions. |
| Screening process | A single reviewer (the first author) assessed eligibility based on each video’s title, description, and a review of its content. |
| Retrieval date | 6/15/26 to 9/15/26 |

Supplemental Table 2. Characteristics of included videos

| Video Type | ID | Title | Length | Link |
|---|---|---|---|---|
| Intake | 1 | Mock counseling "Intake" First session | 10:09 | https://www.youtube.com/watch?v=cawHVBFpOGw |
| | 2 | Scenario 1: Trauma Informed Care Intake Role Play with Dr. Laurie Markoff | 17:27 | https://www.youtube.com/watch?v=RvoPK4kOSeg |
| | 3 | Intake and Assessment Role-Play Part 1 - Referral and Presenting Problems | 8:10 | https://www.youtube.com/watch?v=OPCYGqzf7Us |
| | 4 | PRACTICE 1 CLINICAL INTERVIEW ROLE-PLAY | 19:59 | https://www.youtube.com/watch?v=7zEEzJNeNPE |
| | 5 | Intake demonstration | 31:43 | http://youtube.com/watch?v=EYGbxNyTQ5M |
| | 6 | Social Work Role Play | 11:29 | https://www.youtube.com/watch?v=qeQpo9BdaQ4 |
| | 7 | Psychology Project: Intake Interview | 18:27 | https://www.youtube.com/watch?v=EVc8JQOKs_M |
| | 8 | 5318 Counseling Skills & Techniques-Role Play | 20:20 | https://www.youtube.com/watch?v=ungI3Do5d0o |
| | 9 | Clinical Psychology Role Play | 8:43 | https://www.youtube.com/watch?v=61efTfF3NAA |
| | 10 | Role play between client and social worker Part-1 | 5:01 | https://www.youtube.com/watch?v=gCbc6gkfaQI |
| | 11 | Mock Counseling Session | 9:03 | https://www.youtube.com/watch?v=kVJwI-F_rf0 |
| Motivational Interviewing | 12 | Motivational Interviewing - Good Example - Alan Lyme | 9:23 | https://www.youtube.com/watch?v=67I6g1I7Zao |
| | 13 | Motivational Interviewing A conversation with "Sal" about managing his asthma | 5:12 | https://www.youtube.com/watch?v=-RXy8Li3ZaE |
| | 14 | Motivational Interviewing Role-Play - Precontemplation Stage - Weight Loss | 11:23 | https://www.youtube.com/watch?v=S7RlcWtlzSI |
| | 15 | Motivational Interviewing Role Play | 3:50 | https://www.youtube.com/watch?v=7WjiXluLAq0 |

(Continued on the next page)

Supplemental Table 2. Characteristics of included videos

| | | | | |
|---|---|---|---|---|
| Demonstration of Bad or Unhelpful Behaviors | 16 | Motivational Interviewing: A Bad Example (Presenter: Alan Lyme) | 3:13 | http://youtube.com/watch?v=_VlvanBFkvI |
| | 17 | How NOT to do Motivational Interviewing: A conversation with "Sal" about managing his asthma | 3:31 | https://www.youtube.com/watch?v=kN7T-cmb_l0 |
| | 18 | Therapy Session: Really Bad Counseling Skills 1 | 24:44 | https://www.youtube.com/watch?v=FQOrzyBuAi8 |
| | 19 | Invalidating Therapist - Role Play - Childhood Trauma: The Apologist | 3:55 | https://www.youtube.com/watch?v=HuJIQkJ-_G8 |
| | 20 | Invalidating Therapist - Role Play - Childhood Trauma: The Blank Slate | 3:45 | https://www.youtube.com/watch?v=HuJIQkJ-_G8 |
| | 21 | Invalidating Therapist - Role Play - Childhood Trauma: The Novice Who Took a Training | 4:41 | https://www.youtube.com/watch?v=HuJIQkJ-_G8 |
| | 22 | Basic Counseling Skills: Kim Lampson, PhD: Flawed attending behavior | 2:00 | https://www.youtube.com/watch?v=ankWMLepQsE |
| | 23 | Basic Counseling Skills: Kim Lampson, PhD: Flawed empathy | 2:21 | https://www.youtube.com/watch?v=ankWMLepQsE |
| | 24 | Basic Counseling Skills: Kim Lampson, PhD: Flawed goal setting | 3:40 | https://www.youtube.com/watch?v=ankWMLepQsE |
| | 25 | Worst Counseling Center EVER! | 4:48 | https://www.youtube.com/watch?v=48wzZww2RUk |

Supplemental Table 3. Model and prompt details.

| Provider | Identifier | Access date | Temperature |
|---|---|---|---|
| Google Gemini | gemini-3.1-pro-preview | 9/20/2026 | 1 |
| OpenAI | gpt-6.1-sol | 11/1/2026 | 1 |

Note. gemini-3.1-pro-preview is a preview endpoint that may be modified or discontinued without notice.

VLM prompt:

You are an expert in counseling psychology and nonverbal communication analysis.

Analyze the therapy session video and produce a STRUCTURED JSON transcript of the full conversation, capturing:

1. the spoken dialogue,

2. nonverbal communication of both the therapist and the client, and

3. grounded emotional expression when it is supported by observable cues.

Nonverbal behavior categories to capture — these names are also the allowed values of a cue's "category" field:

- facial_expressions: Movements and expressions of the face indicating affect. Examples: frowning, smiling, neutral affect

- eye_contact: Gaze directed at the other person's face or eyes. Examples: establishing eye contact, sustained contact, breaking contact

- visual_attention: Gaze directed at anything other than the other person. Examples: looking at paperwork, looking at a laptop, looking at the camera, looking down or away

- body_posture: Overall body position and orientation. Examples: open/closed posture, leaning in/away, slouching

- gestures: Movements of hands or body used to communicate. Examples: hand movements, self-touching, pointing

- head_movements: Movements of the head signaling engagement or response. Examples: nodding, shaking, tilting

- interpersonal_distance: Physical distance between therapist and client. Examples: leaning closer, moving away

- vocal_cues: Nonverbal aspects of speech. Examples: tone, pace, pauses, silence

ORDERING AND TURN STRUCTURE

- Emit turns in strict chronological order. "speaker" must be exactly "therapist" or "client".

- Within each turn, list cues in each array in the chronological order in which they occur.

- When a nonverbal behavior occurs without speech, emit a turn whose "utterance" is exactly "(no speech)" and set "anchor_text" to "" on every cue in that turn.

SPLIT LONG TURNS

- Never let a single turn cover more than about 15 seconds of speech.

- When one person speaks continuously for longer than that, break the speech into several consecutive turns for that SAME speaker, each with its own timestamp. Split at natural clause or sentence boundaries, never mid-word.

- Consecutive turns by the same speaker are expected and correct. Do not merge them.

CUE OBJECTS

- ONE BEHAVIOR PER CUE OBJECT. Never combine behaviors in the "text" field of a single cue. Do not join behaviors with commas or semicolons. "looks at therapist, small smile" is WRONG; emit two separate cue objects.

- A turn may contain as many cue objects as the footage warrants. There is no upper limit on cues per utterance; capture every behavior that meets the prioritization criteria below.

Every cue's "type" is exactly one of two values:

- "descriptive": a single, directly observable action or state, visual or audible, that another viewer could confirm or disconfirm by watching the clip. Be specific about the exact behavior, its direction or target when relevant, and its timing relative to speech.

Examples: "single emphatic nod", "closed-lip smile", "gaze shifts to laptop", "open-palmed hand gesture", "quiet voice", "leans forward".

- "interpretive": an inference about emotion, internal state, or interactional function that cannot be strictly confirmed, only judged plausible from the observable evidence. Include the likely interactional function or underlying emotion when visually supported.

Examples: "appears encouraging", "seems tentative", "tone sounds warm".

If a cue could be either, prefer "descriptive" and state the observation concretely. A turn's "cues" array may be empty.

CUE CATEGORY — descriptive cues only

- Every "descriptive" cue carries a "category": exactly one of the category names listed above, naming the channel the behavior physically occurs in.

- "eye_contact" and "visual_attention" are separated by the TARGET of the gaze, never by what the gaze seems to mean. Gaze at the other person's face or eyes is eye_contact. Gaze at anything else — papers, a form, a laptop, the camera, the floor, or simply away — is visual_attention, even when it happens mid-conversation.

- Categorize the behavior itself, not what it means. "nods head" is head_movements even though it signals agreement; "leans forward" is body_posture even though it signals interest.

- If a behavior seems to span two channels, that is a sign it is not atomic: split it into separate cue objects, one per channel. "turns head toward client" (head_movements) and "establishes eye contact" (eye_contact) are two cues, not one.

- Every "interpretive" cue must set "category" to exactly "none". An interpretation is a conclusion drawn from one or more channels rather than a behavior located in any single one, so it is never categorized. Do not guess a category for it.

CUE TIMING — locate each behavior WITHIN the turn

Each cue carries its own "timestamp" and "anchor_text" placing it inside the utterance. A cue's timestamp is when THAT behavior begins, which is usually LATER than the turn's timestamp. Copying the turn's timestamp onto every cue is WRONG.

- "timestamp": MM:SS from the start of the video, at the moment the behavior begins. It must fall between this turn's timestamp and the start of the next turn.

- "anchor_text": copy VERBATIM the few words (roughly 2-6) from this turn's "utterance" that are being spoken as the behavior begins. The string must appear in "utterance" character for character — same spelling, same filler words, same punctuation. Do not paraphrase, do not summarize, do not invent words that are not in the utterance. If a behavior begins before any words are spoken, anchor it to the FIRST words of the utterance. If it begins after the last word, anchor it to the LAST words.

Return ONLY JSON in this shape, with no surrounding prose or code fences:

{"video_title": "<title>",

"turns": [

{"timestamp": "MM:SS",

"speaker": "therapist" | "client",

"utterance": "<spoken words, or \"(no speech)\" for a standalone nonverbal moment>",

"cues": [

{"text": "<one atomic behavior>",

"type": "descriptive" | "interpretive",

"category": "<one of: facial_expressions, eye_contact, visual_attention, body_posture, gestures, head_movements, interpersonal_distance, vocal_cues — or "none" when type is interpretive>", "timestamp": "MM:SS",

"anchor_text": "<verbatim words being spoken as the behavior begins, or "">"}]}]}

LLM prompt:

System message: You are an expert evaluator of therapy sessions.

User message:

You are an expert clinical supervisor trained in the ENACT (ENhancing Assessment of Common Therapeutic factors) framework. Your task is to evaluate a therapy session transcript against the ENACT checklist and produce a structured assessment.

ENACT Checklist:

[Full text of the ENACT checklist inserted here]

Transcript:

[Session transcript inserted here]

Provide a summary of the session context (e.g., session type, client familiarity) and evaluate ENACT competency domains based on the context.

Domain 7 ASSESSMENT OF HARM & DEVELOPING RESPONSE PLAN is removed due to the complexity of assessing harm-related skills. Focus on the other 14 domains for this evaluation.

For each ENACT competency domain, do the following:

1. Check observed behaviors from each tier:

  - Unhelpful/harmful behaviors (if any are present, assign Level 1)

  - Basic helping skills (list which ones were observed vs. absent)

  - Advanced helping skills (list which ones were observed vs. absent)

2. Assign a level by applying these rules in order — the FIRST rule that matches wins:

  a. Level 1: any unhelpful or harmful behavior was observed (true)

  b. null:  more than half of this domain's basic skills are null

    (also set confidence to "not assessable")

  c. Level 4: at least one advanced skill observed, other than

    completes_all_basic_skills, whether or not all basic skills were observed.

  d. Level 3: all assessable basic skills observed, and no advanced skill observed

e. Level 2: some but not all assessable basic skills observed, or none observed, and no advanced skill observed

"Assessable" means non-null. Skills marked null are excluded from the level calculation entirely — never count them against the therapist.

3. Write a brief rationale for your level assignment.

ASSESSABILITY RULES:

1. Do not infer unobserved behaviors.

2. Use exactly one of three values for each behavior:

- true → explicit evidence of the behavior exists in the transcript
- false → the opportunity to demonstrate this skill clearly arose in the session, but the behavior was absent
- null → no meaningful opportunity arose (e.g., topic never came up, session type doesn't require it, or insufficient context to judge)

3. Determining false vs. null — ask this question for each skill:

"Did the session create a natural or necessary opportunity for this skill?"

- YES + skill demonstrated → true
- YES + skill not demonstrated → false
- NO, or unclear → null

Common null scenarios (not exhaustive):

- Introductions/name exchange: null if the client's name was already known, or introductions weren't contextually expected
- Confidentiality briefing: null if this is not a formal intake/first session

4. Domain-level assessability (the level rules in step 2 take precedence):

- If exactly half (or fewer) of a domain's basic skills are null, score only the assessable skills and lower confidence to "medium" or "low" accordingly
- Never assign Level 2 solely because context-dependent skills were absent without opportunity

Return a single JSON object containing all 14 remaining competency domains and strictly following the structure below:

```
{"session_context": {
   "session_number": "1",
   "session_type": "intake",
   "client_known_beforehand": false,
   "notes": "First in-person intake session; no prior relationship established"
 },
 "domains": [
   {"domain_number": 1,
    "domain_name": "Non-Verbal Communication",
    "level": 4,
    "unhelpful_behaviors": {
      "engages_in_other_activities": false,
      "laughs_at_client": false,
      "uses_inappropriate_facial_expressions": false,
      "inappropriate_physical_contact": null
    },
    "basic_skills": {
      "allows_for_silences": true,
      "maintains_appropriate_eye_contact": true,
      "maintains_open_posture": true,
      "continuously_uses_supportive_body_language": true
    },
    "advanced_skills": {
      "completes_all_basic_skills": true,
      "varies_body_language_to_match_client": null
    },
    "confidence": "high",
```

"rationale": "Therapist consistently maintained eye contact and open posture throughout. No physical contact occurred or was warranted during the session",

"evidence": ["00:31 - leans slightly forward", "01:08 - mirrors client's head nod"]

}

"summary": {

"overall_strengths": ["Non-Verbal Communication", "Empathy & Warmth"],

"areas_for_development": ["Explain and Promote Confidentiality"],

"not_assessable_domains": ["Explain and Promote Confidentiality"],

"narrative": "The therapist demonstrated strong foundational skills in rapport and empathy. Several domains could not be assessed due to session context rather than practitioner performance."}}

Supplemental Table 4. Processing time, usage cost, and identified non-verbal behaviors by video.

| Video ID | Length | VLM transcription time (second) | VLM Cost (USD) | LLM evaluation time (second) | LLM cost (USD) | Number of behaviors | Behaviors per min |
|---|---|---|---|---|---|---|---|
| 1 | 10:09 | 262 | 0.58 | 63 | 0.08 | 45 | 4.43 |
| 2 | 17:27 | 123 | 0.47 | 68 | 0.08 | 146 | 8.37 |
| 3 | 8:10 | 133 | 0.40 | 71 | 0.08 | 56 | 6.86 |
| 4 | 19:59 | 178 | 0.62 | 74 | 0.09 | 65 | 3.25 |
| 5 | 31:43 | 193 | 0.86 | 73 | 0.09 | 121 | 3.82 |
| 6 | 11:29 | 166 | 0.51 | 77 | 0.08 | 39 | 3.40 |
| 7 | 18:27 | 150 | 0.54 | 64 | 0.08 | 90 | 4.88 |
| 8 | 20:20 | 217 | 0.73 | 59 | 0.08 | 39 | 1.92 |
| 9 | 8:43 | 110 | 0.34 | 66 | 0.08 | 22 | 2.52 |
| 10 | 5:01 | 61 | 0.19 | 64 | 0.07 | 31 | 6.18 |
| 11 | 9:03 | 116 | 0.36 | 61 | 0.07 | 81 | 8.95 |
| 12 | 9:23 | 140 | 0.44 | 73 | 0.08 | 127 | 13.53 |
| 13 | 5:12 | 86 | 0.25 | 62 | 0.07 | 59 | 11.35 |
| 14 | 11:23 | 125 | 0.42 | 57 | 0.08 | 20 | 1.76 |
| 15 | 3:50 | 90 | 0.25 | 75 | 0.08 | 44 | 11.48 |
| 16 | 3:13 | 88 | 0.23 | 53 | 0.07 | 49 | 15.23 |
| 17 | 3:31 | 74 | 0.20 | 55 | 0.07 | 71 | 20.19 |
| 18 | 24:44 | 141 | 0.59 | 84 | 0.09 | 130 | 5.26 |
| 19 | 3:55 | 101 | 0.28 | 64 | 0.07 | 64 | 16.34 |
| 20 | 3:45 | 79 | 0.22 | 75 | 0.08 | 46 | 12.27 |
| 21 | 4:41 | 87 | 0.25 | 77 | 0.08 | 53 | 11.32 |
| 22 | 2:00 | 60 | 0.15 | 58 | 0.07 | 50 | 25.00 |
| 23 | 2:21 | 75 | 0.20 | 57 | 0.07 | 39 | 16.60 |
| 24 | 3:40 | 76 | 0.22 | 70 | 0.07 | 35 | 9.55 |
| 25 | 4:48 | 116 | 0.32 | 61 | 0.07 | 36 | 7.50 |

Supplemental Table 5. Number of videos without a valid domain level, by domain and rater.

| Domain | Abbreviated Name | LLM | Clinician | | | |
|---|---|---|---|---|---|---|
| | | | #1 | #2 | #3 | #4 |
| 1 | Non-Verbal Comm. | 3 | 2 | 2 | 0 | 5 |
| 2 | Verbal Comm. | 0 | 0 | 0 | 0 | 2 |
| 3 | Confidentiality | 12 | 2 | 7 | 0 | 13 |
| 4 | Rapport Building | 11 | 0 | 0 | 0 | 6 |
| 5 | Normalizing Feelings | 0 | 0 | 0 | 0 | 1 |
| 6 | Empathy & Warmth | 0 | 0 | 0 | 0 | 0 |
| 8 | Social Functioning | 0 | 2 | 0 | 0 | 5 |
| 9 | Client's Explanation | 0 | 2 | 1 | 0 | 6 |
| 10 | Family Involvement | 3 | 1 | 7 | 1 | 10 |
| 11 | Goal Setting | 0 | 1 | 1 | 0 | 1 |
| 12 | Hope for Change | 0 | 0 | 0 | 0 | 8 |
| 13 | Coping Mechanisms | 0 | 0 | 3 | 0 | 8 |
| 14 | Psychoeducation | 3 | 0 | 2 | 0 | 7 |
| 15 | Feedback Elicitation | 2 | 2 | 2 | 0 | 6 |
| | Total | 34 | 12 | 25 | 1 | 78 |

Note. Counts are out of 25 videos. A rating was counted as not valid when the LLM reported it as Not Assessable or the clinician left the domain level blank. LLM=gpt-6.1-sol. Clinician #1–#4 are the four clinician raters. Domain 7 was excluded.

Supplemental Table 6. Between-clinician and AI-clinician ICC with bootstrapped 95% CI for all ENACT domains evaluated.

| Comparison | Domain | Abbreviated Name | Full Name | *N* | ICC(1) | 95% CI |
|---|---|---|---|---|---|---|
| Between Clinician | 1 | Non-Verbal Comm. | Non-Verbal Communication | 25 | .59 | .36-.79 |
| | 2 | Verbal Comm. | Verbal Communication | 25 | .93 | .84-.97 |
| | 3 | Confidentiality | Explain and Promote Confidentiality | 25 | .60 | .31-.83 |
| | 4 | Rapport Building | Rapport Building and Self-Disclosure | 25 | .62 | .43-.80 |
| | 5 | Normalizing Feelings | Exploration & Normalization of Feelings | 25 | .68 | .50-.87 |
| | 6 | Empathy & Warmth | Demonstrate Empathy, Warmth & Genuineness | 25 | .71 | .56-.81 |
| | 8 | Social Functioning | Connect to Social Functioning & Impact on Life | 25 | .63 | .46-.78 |
| | 9 | Client's Explanation | Explore Client's Explanation for Problem | 25 | .73 | .52-.87 |
| | 10 | Family Involvement | Involvement of Family and Significant Others | 24 | .43 | .19-.70 |
| | 11 | Goal Setting | Collaborative Goal Setting | 25 | .77 | .61-.88 |
| | 12 | Hope for Change | Promote Realistic Hope for Change | 25 | .79 | .63-.89 |
| | 13 | Coping Mechanisms | Incorporate Coping Mechanisms & Prior Solutions | 25 | .57 | .36-.76 |
| | 14 | Psychoeducation | Psychoeducation with Local Terminology | 25 | .65 | .45-.80 |
| | 15 | Feedback Elicitation | Elicitation of Feedback | 25 | .65 | .46-.82 |
| Clinician vs. AI | 1 | Non-Verbal Comm. | Non-Verbal Communication | 22 | .57 | .23-.80 |
| | 2 | Verbal Comm. | Verbal Communication | 25 | .55 | .15-.81 |
| | 3 | Confidentiality | Explain and Promote Confidentiality | 13 | -.01 | -.38-.60 |
| | 4 | Rapport Building | Rapport Building and Self-Disclosure | 14 | .60 | .30-.80 |
| | 5 | Normalizing Feelings | Exploration & Normalization of Feelings | 25 | .63 | .26-.84 |
| | 6 | Empathy & Warmth | Demonstrate Empathy, Warmth & Genuineness | 25 | .61 | .27-.78 |
| | 8 | Social Functioning | Connect to Social Functioning & Impact on Life | 25 | .54 | .32-.73 |
| | 9 | Client's Explanation | Explore Client's Explanation for Problem | 25 | .69 | .46-.85 |
| | 10 | Family Involvement | Involvement of Family and Significant Others | 22 | .56 | .32-.75 |
| | 11 | Goal Setting | Collaborative Goal Setting | 25 | .70 | .31-.87 |
| | 12 | Hope for Change | Promote Realistic Hope for Change | 25 | .51 | .20-.74 |
| | 13 | Coping Mechanisms | Incorporate Coping Mechanisms & Prior Solutions | 25 | .60 | .42-.77 |
| | 14 | Psychoeducation | Psychoeducation with Local Terminology | 22 | .46 | .14-.72 |
| | 15 | Feedback Elicitation | Elicitation of Feedback | 23 | .57 | .26-.79 |

Note. *N*=number of videos with a valid rating from at least one pair of raters in the comparison for that domain, i.e., rated by at least two clinicians (between-clinician) or by the AI and at least one clinician (clinician-AI). ICC(1) was estimated on stacked rater pairs. 95% CIs are bias-corrected and accelerated (BCa) bootstrap intervals, resampling videos 2,000 times.
